# Mass Spectrometry-Based Quantification of Orexin Species in Human Cerebrospinal Fluid Reveals Differential Dynamics Associated with Sleep

**DOI:** 10.64898/2025.12.06.25341754

**Authors:** Kanta Horie, Rama K. Koppisetti, Christopher J. Steger, Giuseppe Plazzi, Brendan P. Lucey

**Author notes:** contributed equally (Rama K. Koppisetti, Christopher J. Steger). corresponding author: Brendan P. Lucey, MD, MSCI, Department of Neurology, Washington University School of Medicine, St Louis, MO 63108.

## Abstract

Orexin neuropeptides are central to sleep-wake regulation. However, current cerebrospinal fluid (CSF) assays primarily detect inactive orexin-A fragments using radioimmunoassay (RIA), which limits mechanistic understanding. We developed a high-sensitivity mass spectrometry (MS) assay that can comprehensively quantify human CSF orexin species, including the biologically active long orexin-A and orexin-B, as well as their metabolites and prepro-orexin that are biologically inactive. In participants with narcolepsy type 1 (NT1), all orexin species were significantly reduced compared to those with narcolepsy type 2 (NT2) or idiopathic hypersomnia (IH). MS detected all orexin peptides in NT1 samples below the RIA detection threshold. Notably, the ratio of short to long orexin-B peptides differentiated NT2 from IH, suggesting altered orexin peptide metabolism. Sleep deprivation increased all orexin species with distinct temporal dynamics across peptide forms. The stable isotope labeling kinetics method using MS revealed increased turnover of orexin-A and orexin-B, but not prepro-orexin, during sleep deprivation, indicating activity-dependent regulation. These findings demonstrate that MS enables comprehensive profiling of orexin dynamics and reveals biologically relevant differences in peptide processing and turnover under different sleep conditions. This approach is a powerful tool for advancing our understanding of orexin physiology and its role in sleep-wake pathophysiology.

## INTRODUCTION

The orexin system is involved in regulating multiple physiological functions such as sleep-wake activity, energy metabolism, feeding behavior, spontaneous physical activity, mood and emotional regulation, and reward mechanisms. Orexin-A and orexin-B (also called hypocretin-1 and hypocretin-2) involved in this system are two wake-promoting neuropeptides encoded by a common precursor 131 amino acid polypeptide, prepro-orexin (Tsujino and Sakurai, 2009). Prepro-orexin undergoes post-translational proteolytic cleavage to form orexin-A and orexin-B. Orexin-A is a 33 amino acid residue peptide with a N-terminal glutamine cyclized to pyroglutamate and two intrachain disulfide bonds. Orexin-B is a 28 amino acid residue peptide consisting of two alpha-helices connected with a short linker. The C-termini of both orexin-A and orexin-B are amidated. Human orexin-A and orexin-B have 46% homology with their C-terminal side being well-conserved.

Neurons producing orexin peptides are exclusively localized to the perifornical area and the lateral and posterior hypothalamic area, and project to the brainstem nuclei, amygdala, hippocampus, and cerebral cortex (Date et al., 1999; Elias et al., 1998; Nambu et al., 1999; Peyron et al., 1998). Orexin-A and orexin-B bind to two G protein-coupled receptors, orexin receptor 1 (OX1R) and orexin receptor 2 (OX2R) (Tsujino and Sakurai, 2009). The orexin system regulates sleep-wake activity, feeding behavior, energy homeostasis, and the reward system. Due to loss of 85-95% of orexinergic neurons, individuals with narcolepsy type I (NT1) have cerebrospinal fluid (CSF) immunoreactive concentrations of orexin-A ≤110 pg/ml. The current gold standard for measuring CSF orexin-A is a polyclonal radioimmunoassay (RIA) that differentiates individuals with NT1 from healthy controls, individuals with narcolepsy type 2 (NT2), and idiopathic hypersomnia (IH) (Mignot E, 2002). This RIA measures the N-terminal of orexin-A (Sakai et al., 2019). Although some NT2 patients have been reported to have a partial loss of orexinergic neurons, most patients with NT2 have normal levels of CSF orexin-A. Previous work has shown that the RIA measures <10% of the intact form of orexin-A peptide, suggesting that the RIA for CSF orexin-A primarily measures “unauthentic” short form of orexin-A-related metabolites (Sakai et al., 2019). Importantly, the intact or long forms of orexin-A and orexin-B peptides show the functionalities to bind to orexin-receptors while the short forms do not (Lang et al., 2004). A recent study found that the 1-16 fragment of orexin-A is the likely source of most immunoreactivity detected by the RIA (Vialaret et al., 2025).

Since the functions of the orexin system are commonly disrupted in multiple diseases, its modulation is a potential therapeutic approach for neurodegenerative, metabolic, and addictive disorders. For example, dual orexin receptor antagonists (DORAs), such as suvorexant and lemborexant, are approved to treat insomnia and have been shown in transgenic mice and humans to decrease both amyloid-β (Aβ) and phosphorylated tau in human CSF and AD pathology (Kang et al., 2009; Lucey et al., 2023; Parhizkar et al., 2025).

The full biology of prepro-orexin, orexin-A, and orexin-B is not well-understood and is a major barrier to developing new therapies targeting orexin. Most of the information we know about orexin peptides is based on studies using RIA which majorly detects the short metabolites of orexin-A species in CSF. Orexin-A regulates feeding behavior (Sakurai et al., 1998), sleep-wake activity (Peyron et al., 2000), and reward seeking (Harris et al., 2005). CSF orexin-A levels also fluctuate with a diurnal pattern consistent with a neurotransmitter that regulates sleep-wake activity (Grady et al., 2006; Mignot et al., 2021; Salomon et al., 2003). However, the turnover kinetics of orexin peptides are not known. In particular, the response of production and clearance of these wake-promoting control peptides are important to understand the dynamic control of sleep signals. The role of orexin-B is less well-understood, but there is evidence it differs from orexin-A. For example, orexin-B regulates behavioral and other processes such as thermo-regulation but is less involved in control of energy metabolism or normal food intake (Cai et al., 2001; Ida et al., 1999; Jones et al., 2001; Székely et al., 2002). These differences in regulatory functions of orexin-A and orexin-B may be due to differences in orexin receptor binding. Orexin-A shows similar affinity to both OXR1 and OXR2, while orexin-B shows higher affinity to OXR2 over OXR1 (Mieda et al., 2011).

There are immunoassays for prepro-orexin and orexin-B, but they are less commonly used due to technical limitations in sensitivity, specificity, and standardization. Therefore, orexin peptides remain poorly characterized at the protein structure and isoform level. Mass spectrometry (MS), with its greater analytical specificity, has the potential to identify biological differences between orexin peptides. An immunoassay is less selective than MS and unable to characterize the entire protein. Further, MS is more sensitive than an immunoassay permitting more precise measurement of orexin concentrations. In this study, we report the development and validation of a novel MS assay to measure the concentrations and kinetics of intact prepro-orexin, orexin-A, and orexin-B, and their metabolites in human CSF that will lay the foundation for future studies of the orexin system.

## METHODS

### Participant Characteristics and Sample Collection

CSF previously collected from forty-five participants with NT1 (N=15), NT2 (N=15), and IH (N=15) was used to validate the MS assay. Participants were confirmed to have NT1, NT2, and IH per International Classification of Sleep Disorders criteria by a sleep medicine specialist and multiple sleep latency tests (MSLTs) results. As CSF orexin-A immunoreactivity was known in all samples, all NT2 and IH samples had normal (>110 pg/ml) CSF orexin-A immunoreactivity. Clinical characteristics are reported in Table 1.

**Table 1:** Participant Characteristics.

| Table 1: Participant Characteristics |  |  |  |
| --- | --- | --- | --- |
| Central Disorders of Hypersomnia Participants |  |  |  |
|  | NT1 | NT2 | IH |
| Number (n) | 15 | 15 | 15 |
| Age (years±SD) | 19.9±1.33 | 26.3±1.75 | 33.5±2.24 |
| Sex (male%) | 60% | 53% | 60% |
| Caucasian (%) | 53% | 73% | 67% |
| MSLT Sleep Latency (min) | 2.35±2.82 | 4.10±1.83 | 4.40±1.81 |
| Number of MSLT SOREMP (n±SD) | 3.80±0.68 | 3.53±0.83 | 0.40±0.50 |
| HLA-DQB1*0602 (%) | 100% | 46.7% | 14.3% |
| Orexin-A (pg/ml±SD) | 25.7±1.71 | 330.6±22.04 | 332.5±22.16 |
| % Orexin-A ≤110 pg/ml | 100% | 0% | 0% |
| % Orexin-A ≤200 pg/ml | 100% | 0% | 0% |
| Lumbar Catheter Participants |  |  |  |
|  | Control | Sleep-deprived |  |
| Number (n) | 4 | 4 |  |
| Age (years±SD) | 48.3±11.75 | 48.1±11.89 |  |
| Sex (male%) | 50% | 50% |  |
| Caucasian (%) | 25% | 25% |  |
| Overnight Sleep Time (minutes, mean±SD) | 394±44.16 | 34.88±25.55 |  |
| Sleep Efficiency (% , mean±SD) | 72.85%±11.13 | 6.43%±4.65 |  |
| NT1: narcolepsy type 1; NT2: narcolepsy type 2; IH: idiopathic hypersomnia; SD: standard deviation; pg: picogram; ml: milliliters; HLA: Human Leukocyte Antigen; PSG: nocturnal polysomnography; MSLT: Multiple Sleep Latency Test; SOREMP: sleep onset rapid eye movement (REM) period, defined as REM sleep occurring ≤15 minutes following sleep onset. Note: Four participants in the lumbar catheter study completed both the normal sleep control and sleep-deprived conditions. |  |  |  |

Serial CSF samples collected from four participants aged 30 to 60 years who completed both sleep deprivation and normal sleep control conditions in a previously published study (Lucey et al., 2018) were analyzed. Participants completed both behavioral sleep deprivation for 36 hours and a normal sleep control intervention. All participants were in good general health and had no clinical sleep or neurological disorders. All participants were screened to exclude sleep-disordered breathing with a home sleep apnea test and sleep was monitored via polysomnography throughout the CSF collection period. Participant characteristics are shown in Table 1. After an acclimation night, an intrathecal lumbar catheter was placed and collection of CSF was started at 07:00. All participants were kept awake from the time of lumbar catheter placement until the start of the intervention (21:00). At 21:00, control participants were permitted to sleep while the sleep-deprived group was kept awake by nursing staff and did not receive stimulants. Six milliliters of CSF was obtained every 2 hours for 36 hours. Each participant was administered a stable isotope amino acid tracer, [U-^13^C_6_]leucine, starting at 21:00 and infused over 9 hours as previously described (Lucey et al., 2018; Patterson et al., 2015). Tracer:tracee ratios were normalized to plasma leucine concentrations. The lumbar catheter was removed on day 2 at 19:00. Meals were served at 09:00, 13:00, and 18:00.

The study protocol was approved by the Washington University Institutional Review Board.

### Radioimmunoassay for orexin-A

OXA was quantified in duplicate for each CSF sample using a standard validated direct I^125^ RIA that used rabbit polyclonal anti-orexin-A (Phoenix Pharmaceuticals) as previously reported (Mignot E, 2002).

### Sample preparation for mass spectrometry analysis

To purify the samples for MS analysis, the immunoprecipitation (IP) for each orexin species was conducted. For the first IPs of the intact long-form OXA and OXB species, anti-OXA mouse monoclonal antibody (recognizing the C-terminal side of OXA (FUJIFILM Wako) was conjugated with CNBr-activated Sepharose 4B beads (Cytiva) as previously reported (Sato et al., 2018). Since both OXA and OXB have the same amino acid sequence at the C-terminal end, the anti-OXA Wako antibody could capture both species. For the second IP of the metabolite forms of OXA and OXB, anti-OXA mouse monoclonal antibody (recognizing the N-terminal side of OXA (R&D Systems, MAB763) and anti-OXB mouse monoclonal antibody (recognizing the N-terminal side of OXB (R&D Systems, MAB734) were conjugated with the same manner, respectively. For the IP of prepro-orexin, anti-prepro-orexin mouse monoclonal antibody (Sigma, MABN1526) was conjugated on the same beads format, and used for the 2^nd^ IP process together with the OXA metabolite form analysis.

For the first IP, 200 μL of CSF, 10 μL of 20 ng/mL isotope-labelled orexin-A (for OXA1) or orexin-B (for OXB1), 12.5 μL of a slurry consisting of 10% (v/v) IGEPAL CA-630 (Sigma, I8896-100ML), 50 mM Guanidine, and 10x protease inhibitors (Roche, 11836145001), and 20 μL of a 50% slurry of Sepharose beads conjugated with Wako anti-OXA antibody were combined and then incubated for at least 16 hours at 4C. For the second orexin-A (OXA2) and orexin-B (OXB2) IPs, 200 μL of each supernatant from OXA1 (for OXA2) or OXB1 (for OXB2), 10 μL of 50 ng/mL isotope-labelled orexin-A and 50 ng/mL isotope-labelled prepro-orexin (for OXA2) or 50 ng/mL isotope-labelled orexin-B (for OXB2), and 20 μL of a 50% slurry of Sepharose beads conjugated with R&D Systems anti-OXA antibody and 20 μL of a 50% slurry of Sepharose beads conjugated with anti prepro-orexin antibody (for OXA2) or 20 μL of a 50% slurry of Sepharose beads conjugated with R&D Systems anti-OXB antibody were incubated for 2 hours at room temperature (RT). All samples were washed with 1 mL of 25 mM TEABC (total three times). Then, OXA1+2 samples were reduced by the addition of 20 μL of 5 mM dithiothreitol in 25 mM Triethylammonium bicarbonate (TEABC), followed by an incubation at 65 °C for 30 minutes at 1000 rpm on an Eppendorf ThermoMixer C. Then, alkylation was followed with the addition of 20 μL of 10 mM iodoacetamide in 25 mM TEABC, followed by an incubation at RT for 30 minutes in the dark. Then, on-bead digestion was commenced for all samples with the addition of 40 μL of 10 ug/mL trypsin resuspended in 25 mM TEABC, and incubated for 18 hours at 37 °C. The digested peptides were then desalted via reversed-phase solid-phase extraction, using an HLB μElution Plate (Waters, 186001828BA). The reversed-phase sorbent was pre-washed with 200 μL methanol, conditioned with an organic solvent, 200 μL 60% acetonitrile (ACN) with 0.1% formic acid (FA), and re-equilibrated with two additions of an aqueous solvent, 200 μL water with 0.1% FA. Then, the samples were loaded onto the sorbent, washed with two additions of the aqueous solvent, 200 μL water with 0.1% FA, and eluted with the organic solvent, 100 μL 60% ACN with 0.1% FA. The eluent solvent was evaporated *in vacuo*, and the resulting dried peptides were resuspended with 27.5 μL of 3% ACN + 3% FA (for OXA1+2) and 2% ACN + 2% FA (for OXB1+2). All samples were centrifuged at 20,000 g for 10 minutes at 4°C using an angled rotor. Then, 25 μL of sample supernatant was transferred to an MS vial without disturbing the pellet. IP protocols for OXA together with prepro-orexin and OXB are described in Figure 1. The amino acid sequences of the monitored peptides and the schematic of the target region of each antibody is described in Supplemental Figure 1.

**Figure 1:**
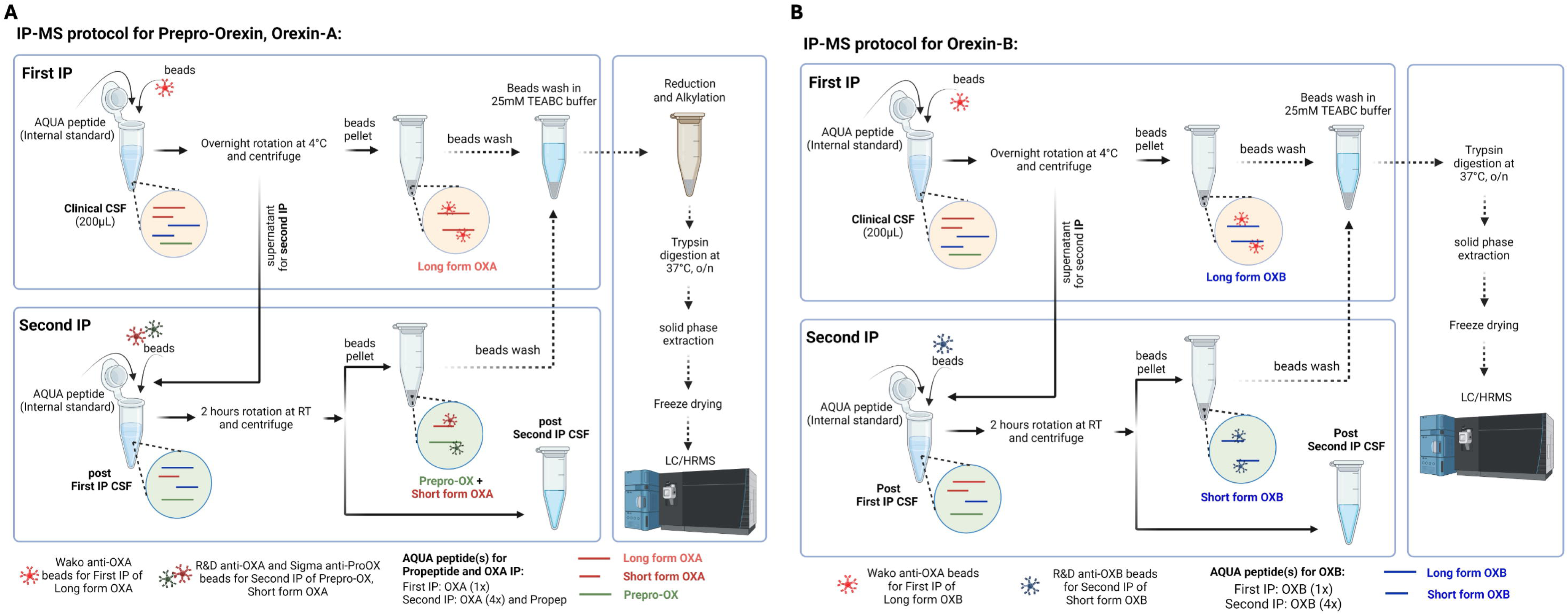
Diagram of the immunopreciptation (IP) mass spectroscopy (MS) protocol. A. Orexin-A and Prepro-orexin. B. Orexin-B.

### Liquid-Chromatography-Mass Spectrometry (LC-MS)

All experiments were performed using a liquid chromatography (Waters ACQUITY UPLC, 176816002)-mass spectrometer (Thermo Orbitrap Eclipse Tribrid, FSN04-10000) equipped with a nanospray ion source. 4.5 µL of sample was auto loaded into a C18 column (Waters, HSS T3 75 µm × 100 µm, 1.8 µm, 186008006), which is heated at 65 °C. The aqueous solvent (mobile phase A) consists of water with 0.1% formic acid (FA), and the organic phase (mobile phase B) consists of an acetonitrile (ACN) with 0.1% FA. The sample loading and precolumn washing was conducted at 100% of solvent A. The peptides were eluted from the C18 column with a 7.7 minute gradient ranging from 0.5 to 4% solvent B, followed by 17.3 minutes gradient from 4 to 40% solvent B, followed by 2 minutes ramp up from 40 to 95% solvent B at a constant flow rate of 0.4 µL/minutes. Finally, the column was washed and equilibrated for another 2 minutes. The eluted peptide ions sprayed from a 10 μm SilicaTip emitter (New Objective, Woburn, MA, USA) into the ion source (spray voltage = 2200 V), were targeted and isolated in the quadrupole. Isolated ions were fragmented by high-energy collisional dissociation (HCD), and ion fragments were detected in the Orbitrap with a resolution of 60,000 for endogenous orexin peptides and 30,000 for internal standard reference peptides at mass range 150–1,300 m/z). Custom automatic gain control was used to prevent overfilling of the orbitrap in MS^2^ mode. The PRM (parallel reaction monitoring) acquired raw data was processed and visualized by using skyline 23.1v (MacCoss Lab, Department of Genome Sciences, University of Washington). Peak areas are extracted for the most abundant MS/MS fragment ions and normalized with internal standard reference peptides.

### Statistics

Participant demographics, DQB1*0602, orexin-A concentrations, and sleep variables were summarized with the mean and standard deviation for continuous variables or percentages for categorical variables. One-way analysis of variance (ANOVA) was performed in GraphPad Prism version 10 for MacOS (Graphpad Software, San Diego, CA, USA) was used to compare group differences in orexin peptide concentrations across hypersomnias of central origin. To conduct cosinor analysis, orexin peptide concentrations were transformed to percent of mean. Orexin peptide concentrations at each time point were expressed as the percent of the mean for each individual participant. Cosinor analysis was then performed in GraphPad Prism version 10 for MacOS as previously described (Lucey et al., 2015) with the linear rise and cosine transformation fitted simultaneously. The cosine transformation was applied to the time variable using 24 hours as the default circadian period. The amplitude (distance between the peak and mesor), acrophase (the time corresponding to the peak of the curve), and the linear rise (change per hour) were calculated for the normal sleep control and sleep-deprived group. All serial CSF orexin data was analyzed using SPSS version 31 (IBM, Armonk, NY) as previously described (Lucey et al., 2018) with general linear mixed models to account for the dependencies among the longitudinal measurements. Intervention group, time of day, and an intervention group x time of day interaction were treated as fixed effects. Random intercepts and slopes for time were used to accommodate individual variation. The significance level was set at p < 0.05 for all statistical analyses and analyses were corrected with Tukey’s multiple comparisons test.

## RESULTS

### Orexin Peptides Differentiate NT1 from NT2 and IH

Participants diagnosed with NT1, NT2, and IH had orexin peptides measured by RIA and IP/MS. CSF orexin-A measured by RIA differentiated NT1 from NT2 and IH (Figure 2). However, 40% (6/15) of participants with NT1 had CSF OXA below the limit of quantification using the RIA. Consistent with the expected finding of a pan-orexin deficiency in NT1, we found that all orexin peptides measured by MS were decreased in NT1 compared to NT2 and IH (Figure 3). Notably, all orexin peptides measured by MS, even from NT1 CSF samples, showed quantifiable MS signals. Further, all orexin-A peptides measured by MS showed a significant linear correlation with orexin-A measured by RIA. As previously reported (Vialaret et al., 2025), the short orexin-A peptide with intact N-terminal region most closely approximates the orexin-A peptide captured by the RIA and showed the greatest correlation compared to the long orexin-A peptide with intact N-terminal region or the long-orexin-A with intact C-terminal region (Figure 4).

**Figure 2:**
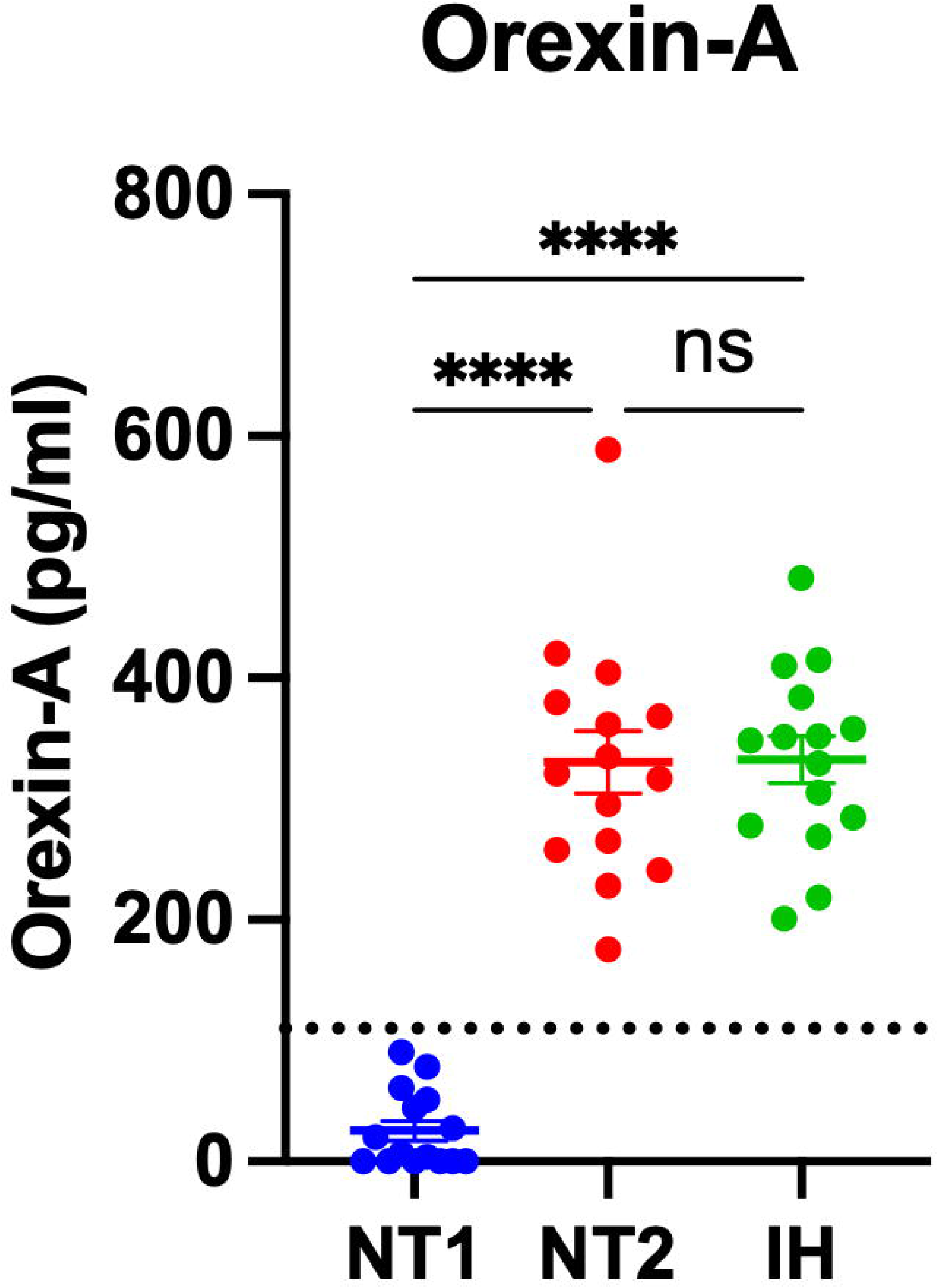
Differentiating narcolepsy type 1 from other central disorders of hypersomnolence by orexin-A measured by radioimmunoassay in human cerebrospinal fluid. Orexin-A was measured by radioimmunoassay (RIA) in the cerebrospinal fluid (CSF) from individuals with narcolepsy type 1 (NT1), narcolepsy type 2 (NT2), and idiopathic hypersomnia (IH). Individuals with NT1 have significantly lower concentrations of CSF orexin-A compared to NT2 and IH. ****p<0.0001 after Tukey correction for multiple comparisons. Dotted line indicates cutoff of 110 pg/ml.

**Figure 3:**
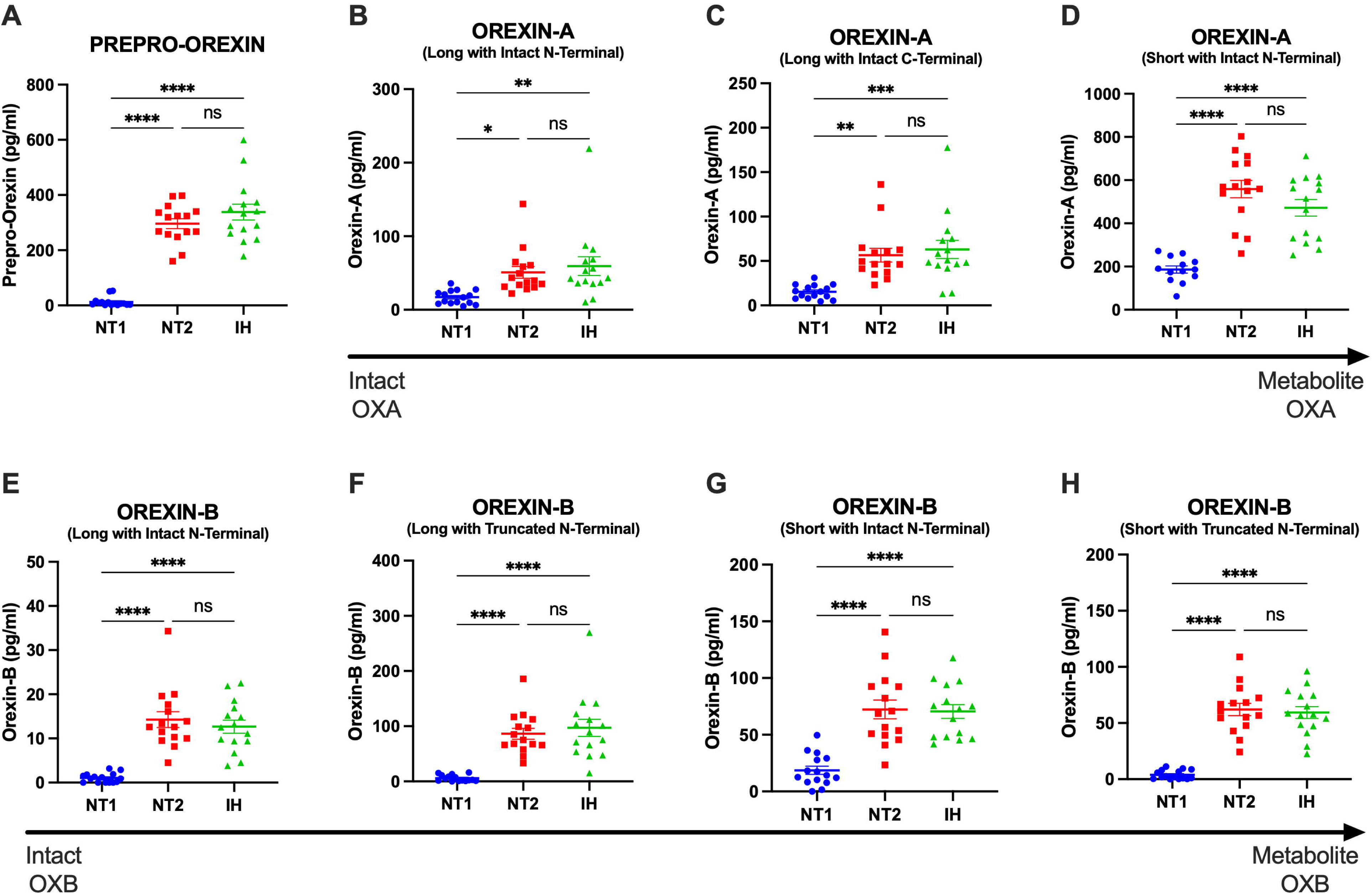
Differentiating narcolepsy type 1 from other central disorders of hypersomnolence by orexin peptides measured by mass spectrometry in human cerebrospinal fluid. Orexin peptides were measured by mass spectrometry (MS) in the cerebrospinal fluid (CSF) from individuals with narcolepsy type 1 (NT1), narcolepsy type 2 (NT2), and idiopathic hypersomnia (IH). A. Prepro-orexin. Individuals with NT1 have significantly lower concentrations of CSF prepro-orexin compared to NT2 and IH. B-D. Orexin-A. Individuals with NT1 have significantly lower concentrations of CSF orexin-A compared to NT2 and IH. E-H. Orexin-B. Individuals with NT1 have significantly lower concentrations of CSF orexin-B compared to NT2 and IH. *p<0.05; **p<0.01; ***p<0.001; ****p<0.0001 after Tukey correction for multiple comparisons.

**Figure 4:**
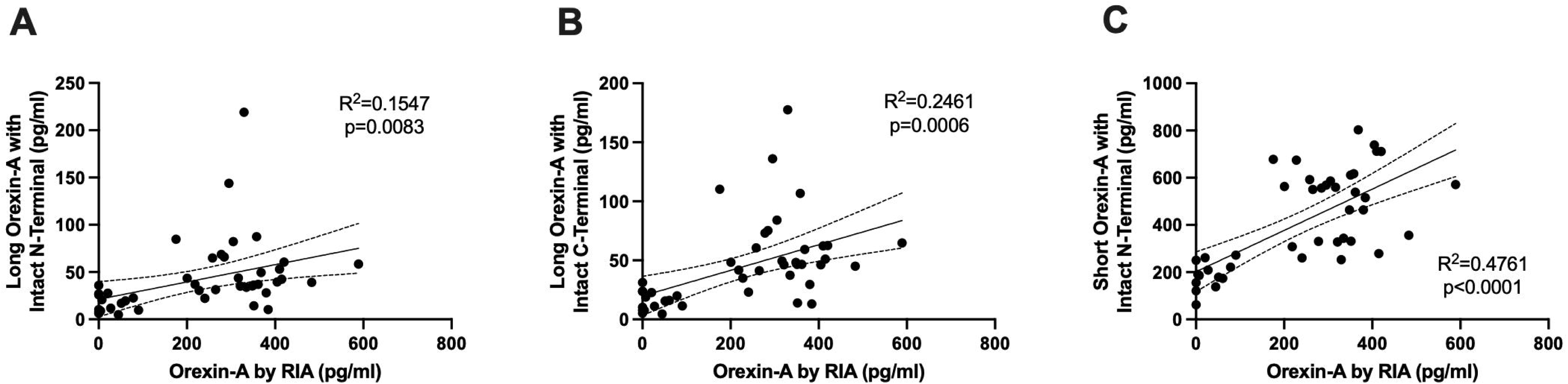
Correlation of orexin-A peptides. Orexin-A measured by radioimmunoassay showed significant linear correlations with the three orexin-A peptides measured by mass spectrometry: Long orexin-A peptide with intact N-terminal (A); Long orexin-A with intact C-terminal (B); Short orexin-A with intact N-terminal (C).

### Ratio of Orexin-B Peptides Differs Between NT2 and IH

Orexin-B maintains stable sleep-wake physiology through its action on OX2R, the receptor most strongly implicated in cataplexy and promoting wakefulness (Willie et al., 2003). While both orexin-A and orexin-B regulate arousal, orexin-B demonstrates higher selectivity for OX2R. OX2R signaling is essential for sustaining consolidated periods of wakefulness and suppressing REM sleep intrusions. Although orexin-B did not differ between NT1, NT2, and IH, MS-based analysis can measure specific orexin peptide species which can allow for generating peptide ratios and therefore allows significant advantage in understanding subtle sequence or structural differences in complex biological samples. Therefore, we tested for group differences in the ratio of orexin-B peptides. The ratio of the shortest form of orexin-B over long orexin-B species was significantly decreased in NT2 compared to IH (Figure 5). These findings suggest that the processing of orexin-B could underlie the pathophysiology of NT2 or reflect disrupted REM sleep in non-NT1 hypersomnias.

**Figure 5:**
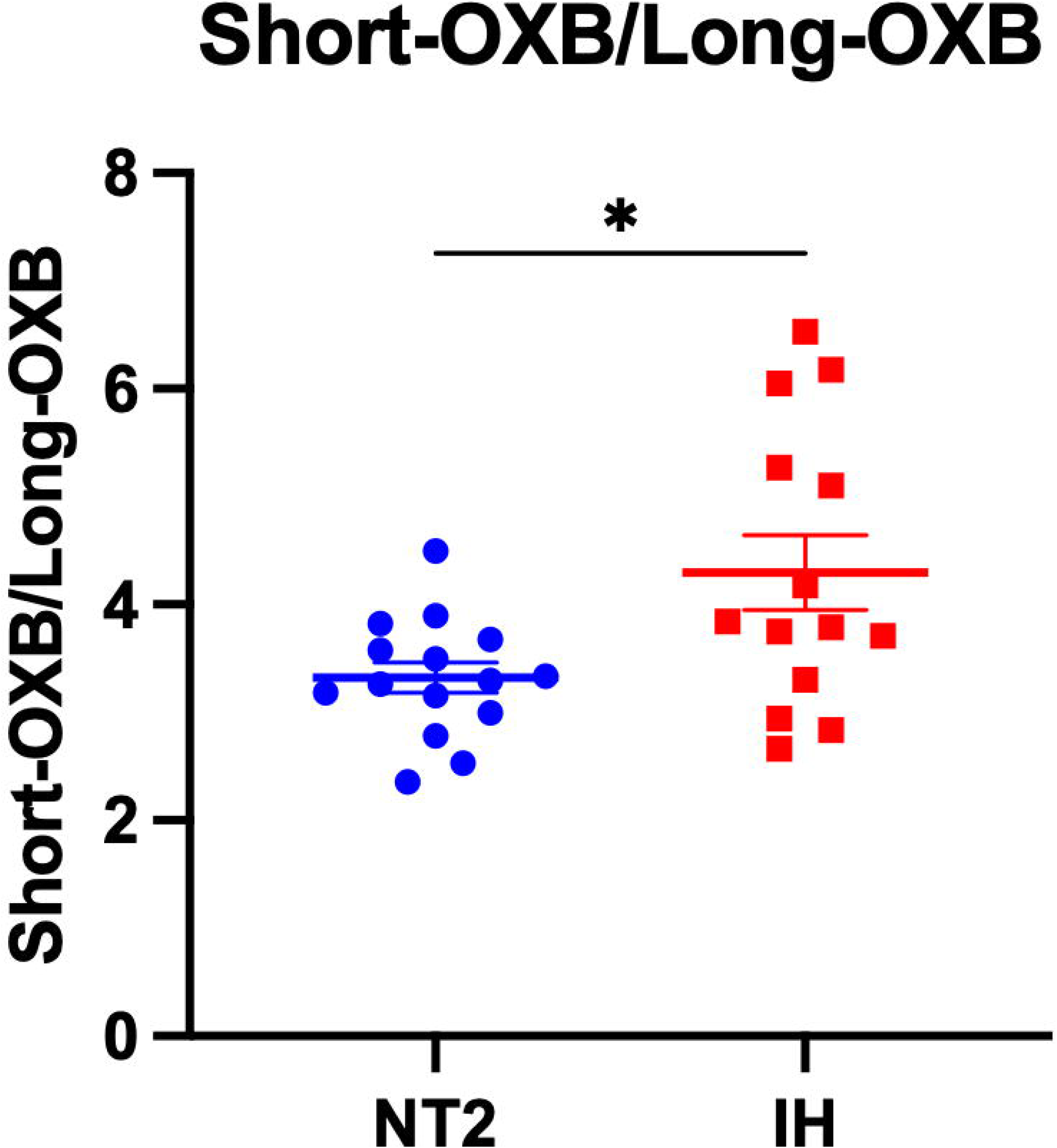
Differentiating narcolepsy type 2 and idiopathic hypersomnia. Orexin-B (OXB) peptides were measured by mass spectrometry in the cerebrospinal fluid (CSF) of individuals with narcolepsy type 2 (NT2) and idiopathic hypersomnia (IH). The ratio of the short form of orexin-B/long form of orexin-B significantly differentiated NT2 from IH. *p<0.05.

### Effect of Sleep Deprivation on CSF Orexin Peptides

Prior work in humans found that lumbar CSF orexin-A immunoreactivity fluctuates ∼10% across the 24-hour day (Salomon et al., 2003). In CSF samples previously collected every 2 hours over 36 hours from healthy participants under sleep-deprived and normal sleep control conditions, we assessed the diurnal concentration changes of different orexin peptides (Supplemental Figure 2; Supplemental Table 1). After normalization of orexin concentrations to percent of the mean, CSF orexin-A measured by RIA showed a higher amplitude than previously reported (average 22-28%) and without significant differences between sleep-deprived and control groups (Table 2). However, CSF orexin-A measured by RIA was higher at hour 26 in the sleep-deprived group compared to control (Supplemental Figure 3; Supplemental Table 2). Other orexin peptide amplitudes ranged from a low of 7.49% (prepro-orexin in the control group) to a high of 38.68% (long orexin-A peptide with intact N-terminal). In general, the amplitude of CSF orexin peptides was higher in the control group but this was only significant for the long orexin-A peptide with intact N-terminal (Table 2). The acrophase was also not significantly different between groups. Finally, we have previously shown a linear rise in CSF amyloid-β (Aβ) collected from indwelling lumbar catheters (Lucey et al., 2015; Lucey et al., 2017). No consistent direction of linear rise was observed, but there were significant differences between the sleep-deprived and control conditions for prepro-orexin and the short orexin-B peptide (Table 2).

**Table 2:** Amplitude, Acrophase, and Linear Rise of Orexin Peptides.

| Table 2: Amplitude, Acrophase, and Linear Rise of Orexin Peptides |  |  |  |  |  |
| --- | --- | --- | --- | --- | --- |
| Cosinor Measures | Control |  | Sleep-Deprived |  | Group Differences |
|  | Average | SD | Average | SD |  |
| *RIA: Orexin-A* |  |  |  |  |  |
| Amplitude (%) | 28.42 | 10.70 | 22.74 | 13.59 | 0.574 |
| Acrophase (time of day) | 16:33 | 11:01 | 11:59 | 10:17 | 0.651 |
| Linear Rise (%/hour) | -0.59 | 0.86 | 1.29 | 1.77 | 0.115 |
| *MS: Prepro-Orexin* |  |  |  |  |  |
| Amplitude (%) | 7.49 | 5.90 | 9.12 | 3.93 | 0.704 |
| Acrophase (time of day) | 10:09 | 6:22 | 10:24 | 9:22 | 0.976 |
| Linear Rise (%/hour) | -0.59 | 0.54 | 0.30 | 0.48 | **0.034** |
| *MS: Long Orexin-A with Intact N-Terminal* |  |  |  |  |  |
| Amplitude (%) | 38.68 | 12.23 | 23.69 | 9.91 | **0.013** |
| Acrophase (time of day) | 19:20 | 1:41 | 20:54 | 2:23 | 0.448 |
| Linear Rise (%/hour) | -0.34 | 0.42 | 0.38 | 0.26 | 0.123 |
| *MS: Short Orexin-A with Intact N-Terminal* |  |  |  |  |  |
| Amplitude (%) | 15.64 | 10.47 | 9.89 | 8.02 | 0.434 |
| Acrophase (time of day) | 14:15 | 10:14 | 6:39 | 9:19 | 0.162 |
| Linear Rise (%/hour) | -0.42 | 0.72 | 0.80 | 0.53 | 0.117 |
| *MS: Long Orexin-B with Truncated N-Terminal* |  |  |  |  |  |
| Amplitude (%) | 33.94 | 12.95 | 24.14 | 4.44 | 0.322 |
| Acrophase (time of day) | 18:48 | 1:22 | 18:38 | 1:29 | 0.909 |
| Linear Rise (%/hour) | -0.69 | 1.48 | 0.05 | 1.42 | 0.474 |
| *MS: Short Orexin-B with Truncated N-Terminal* |  |  |  |  |  |
| Amplitude (%) | 17.30 | 9.31 | 15.72 | 5.60 | 0.493 |
| Acrophase (time of day) | 18:32 | 1:22 | 16:07 | 10:26 | 0.687 |
| Linear Rise (%/hour) | -0.61 | 0.41 | 0.25 | 0.30 | **0.038** |
| SD: standard deviation; RIA: radioimmunoassay; MS: mass spectrometry |  |  |  |  |  |

To assess for differences in orexin peptide concentrations between sleep-deprived and normal sleep conditions, each orexin peptide concentration was normalized to a baseline for each subject (average of hours 07:00-19:00) before each group’s intervention. This normalization method follows the previously established method as outlined in examining group differences in Aβ, tau, phosphorylated tau, and neuronal pentraxin 2 (Barthélemy et al., 2020; Liu et al., 2023; Lucey et al., 2018; Xiao et al., 2021). All orexin peptides were significantly increased in the sleep-deprived group during the sleep period (Figure 6; Table 3). However, the time courses for different orexin peptides showed variable changes in response to sleep deprivation. Orexin-A measured by RIA was only increased in the latter part of the sleep period (09:00-11:00). Prepro-orexin was elevated during sleep deprivation, but this may be due to differences in the starting baseline concentrations (Supplemental Figure 2). Short orexin-A with intact N-terminal region, which most closely approximates the orexin peptide measured by RIA, showed a bimodal distribution with increases in both the early and late sleep period. Long orexin-A with intact N-terminal, long orexin-B with truncated N-terminal, and short orexin-B with truncated N-terminal were all more consistently increased during sleep deprivation across the sleep period while the control condition showed the clear diurnal rhythm with the increase and decrease of orexin species in CSF.

**Figure 6:**
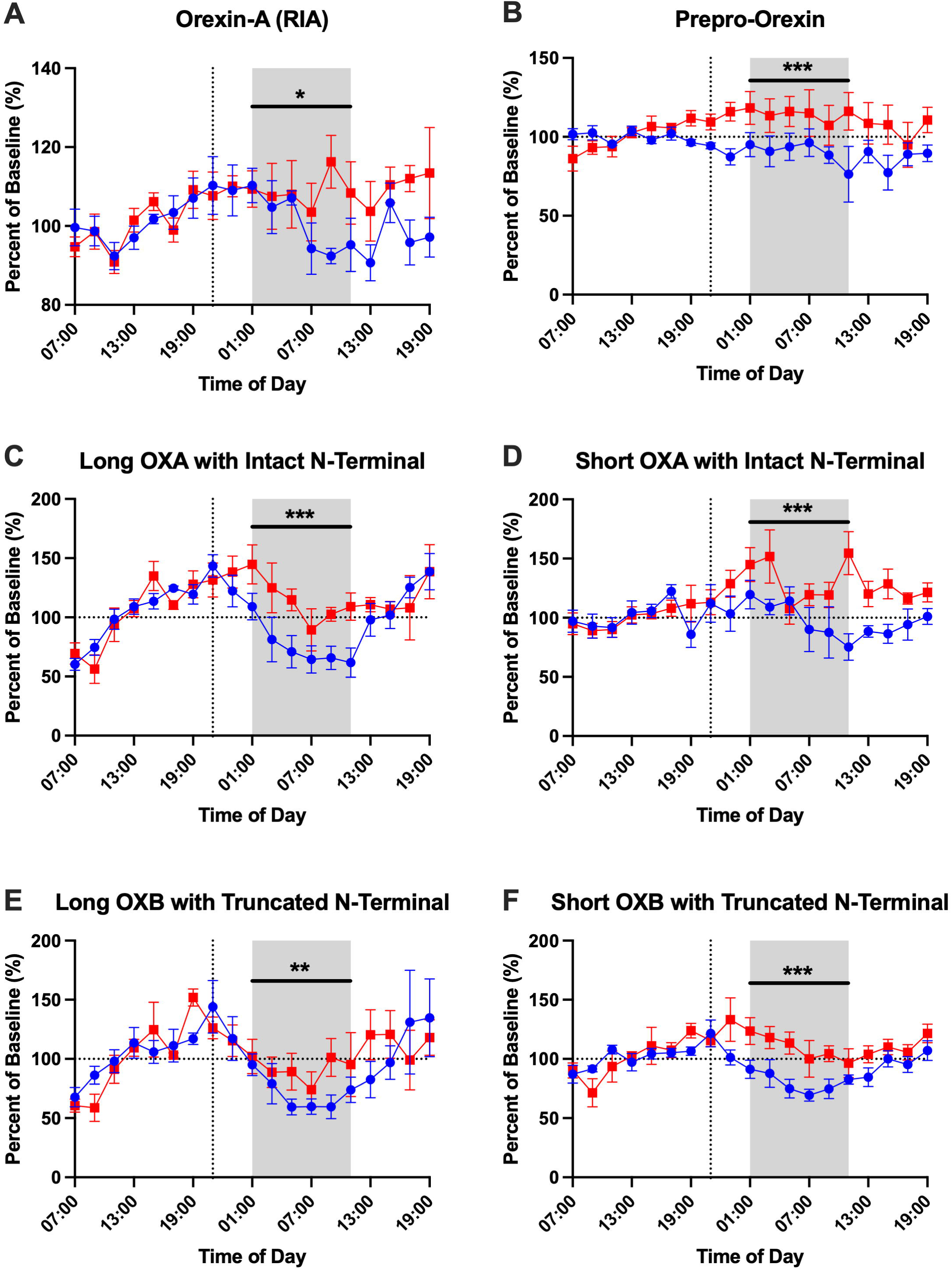
Effect of overnight sleep deprivation on orexin peptide concentrations in cerebrospinal fluid normalized to percent of baseline. Four participants completed both the control normal sleep and sleep-deprived intervention groups. All orexin peptides are normalized to the percent of hours 0-12 (07:00-19:00) baseline. The overnight period during the intervention night was defined as hours 18 to 28 (01:00-11:00) to account for transit time of cerebrospinal fluid (CSF) from the brain to the lumbar catheter (shaded area). Blue: control; Red: sleep-deprived. Error bars indicate standard error. The vertical dashed line is the intervention start time and when ^13^C_6_-leucine was infused. The horizontal dashed line is at 100% of baseline. OXA: orexin-A. OXB: orexin-B. *p<0.05; **p<0.01; ***p<0.001.

**Table 3:**
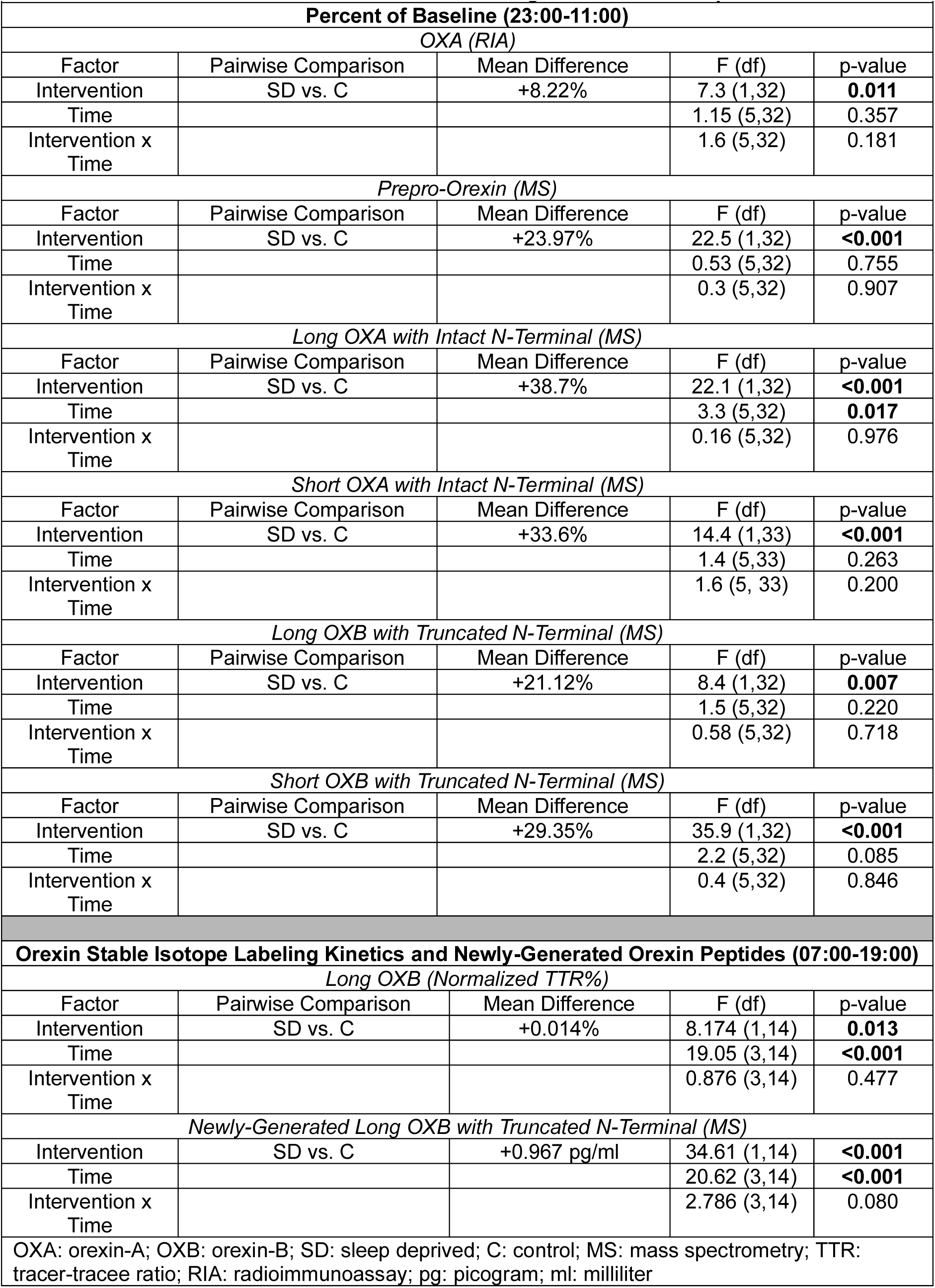
Mixed Model Results for Longitudinal Orexin Peptides.

| **Table 3: Mixed Model Results for Longitudinal Orexin Peptides** |  |  |  |  |
| --- | --- | --- | --- | --- |
| **Percent of Baseline (23:00-11:00)** |  |  |  |  |
| *OXA (RIA)* |  |  |  |  |
| Factor | Pairwise Comparison | Mean Difference | F (df) | p-value |
| Intervention | SD vs. C | +8.22% | 7.3 (1,32) | **0.011** |
| Time |  |  | 1.15 (5,32) | 0.357 |
| Intervention x Time |  |  | 1.6 (5,32) | 0.181 |
| *Prepro-Orexin (MS)* |  |  |  |  |
| Factor | Pairwise Comparison | Mean Difference | F (df) | p-value |
| Intervention | SD vs. C | +23.97% | 22.5 (1,32) | **<0.001** |
| Time |  |  | 0.53 (5,32) | 0.755 |
| Intervention x Time |  |  | 0.3 (5,32) | 0.907 |
| *Long OXA with Intact N-Terminal (MS)* |  |  |  |  |
| Factor | Pairwise Comparison | Mean Difference | F (df) | p-value |
| Intervention | SD vs. C | +38.7% | 22.1 (1,32) | **<0.001** |
| Time |  |  | 3.3 (5,32) | **0.017** |
| Intervention x Time |  |  | 0.16 (5,32) | 0.976 |
| *Short OXA with Intact N-Terminal (MS)* |  |  |  |  |
| Factor | Pairwise Comparison | Mean Difference | F (df) | p-value |
| Intervention | SD vs. C | +33.6% | 14.4 (1,33) | **<0.001** |
| Time |  |  | 1.4 (5,33) | 0.263 |
| Intervention x Time |  |  | 1.6 (5, 33) | 0.200 |
| *Long OXB with Truncated N-Terminal (MS)* |  |  |  |  |
| Factor | Pairwise Comparison | Mean Difference | F (df) | p-value |
| Intervention | SD vs. C | +21.12% | 8.4 (1,32) | **0.007** |
| Time |  |  | 1.5 (5,32) | 0.220 |
| Intervention x Time |  |  | 0.58 (5,32) | 0.718 |
| *Short OXB with Truncated N-Terminal (MS)* |  |  |  |  |
| Factor | Pairwise Comparison | Mean Difference | F (df) | p-value |
| Intervention | SD vs. C | +29.35% | 35.9 (1,32) | **<0.001** |
| Time |  |  | 2.2 (5,32) | 0.085 |
| Intervention x Time |  |  | 0.4 (5,32) | 0.846 |
| **Orexin Stable Isotope Labeling Kinetics and Newly-Generated Orexin Peptides (07:00-19:00)** |  |  |  |  |
| *Long OXB (Normalized TTR%)* |  |  |  |  |
| Factor | Pairwise Comparison | Mean Difference | F (df) | p-value |
| Intervention | SD vs. C | +0.014% | 8.174 (1,14) | **0.013** |
| Time |  |  | 19.05 (3,14) | **<0.001** |
| Intervention x Time |  |  | 0.876 (3,14) | 0.477 |
| *Newly-Generated Long OXB with Truncated N-Terminal (MS)* |  |  |  |  |
| Intervention | SD vs. C | +0.967 pg/ml | 34.61 (1,14) | **<0.001** |
| Time |  |  | 20.62 (3,14) | **<0.001** |
| Intervention x Time |  |  | 2.786 (3,14) | 0.080 |
| OXA: orexin-A; OXB: orexin-B; SD: sleep deprived; C: control; MS: mass spectrometry; TTR: tracer-tracee ratio; RIA: radioimmunoassay; pg: picogram; ml: milliliter |  |  |  |  |

### Orexin Stable Isotope Labeling Kinetics

Prepro-orexin, long orexin-A with intact N-terminal, and long orexin-B with truncated N-terminal were quantified for ^13^C_6_-leucine isotopic enrichment in 3 participants who underwent longitudinal CSF sampling under both sleep-deprived and normal sleep control conditions. Labeled short orexin-A with intact N-terminal and short orexin-B with truncated N-terminal peptides were not quantified. No orexin peptides were quantifiable for a fourth participant with longitudinal CSF samples under the two conditions. The time courses for ^13^C_6_-leucine isotopic enrichment of orexin peptides showed differences between these three labeled orexin peptides (Figure 7). Labeled orexin peptides were not measured in CSF until 5 hours after ^13^C_6_-leucine infusion. This finding is consistent with prior observations of labeled CSF Aβ dynamics in human CSF and most likely represents the transit time from the brain to the lumbar region (Huang et al., 2012; Lucey et al., 2018; Patterson et al., 2015). Prepro-orexin had the highest normalized tracer:tracee ratio (TTR) (∼0.23) followed by long orexin-B with truncated N-terminal (∼0.07). Long orexin-A with intact N-terminal had the lowest normalized TTR (∼0.02). Labeled prepro-orexin showed overlapping labeling curves for both the sleep-deprived and control groups suggesting no group difference in protein turnover. In contrast, the labeled orexin-A and orexin-B peptides increased faster and peaked higher in the sleep-deprived group, but the difference in labeled orexin peptides was only significant for orexin-B. Newly-generated orexin peptides, calculated by TTR x concentration as previously described (Bateman et al., 2009), showed similar relationships (Figure 7; Table 3; Supplemental Table 3). These findings support the hypothesis that prepro-orexin turnover is not affected by changes in the sleep-wake activity, while the turnover of orexin-B is increased during sleep deprivation. These exploratory findings suggest that orexin peptides labeled with stable isotopes are measurable in human CSF and may be responsive to changes in sleep-wake activity.

**Figure 7:**
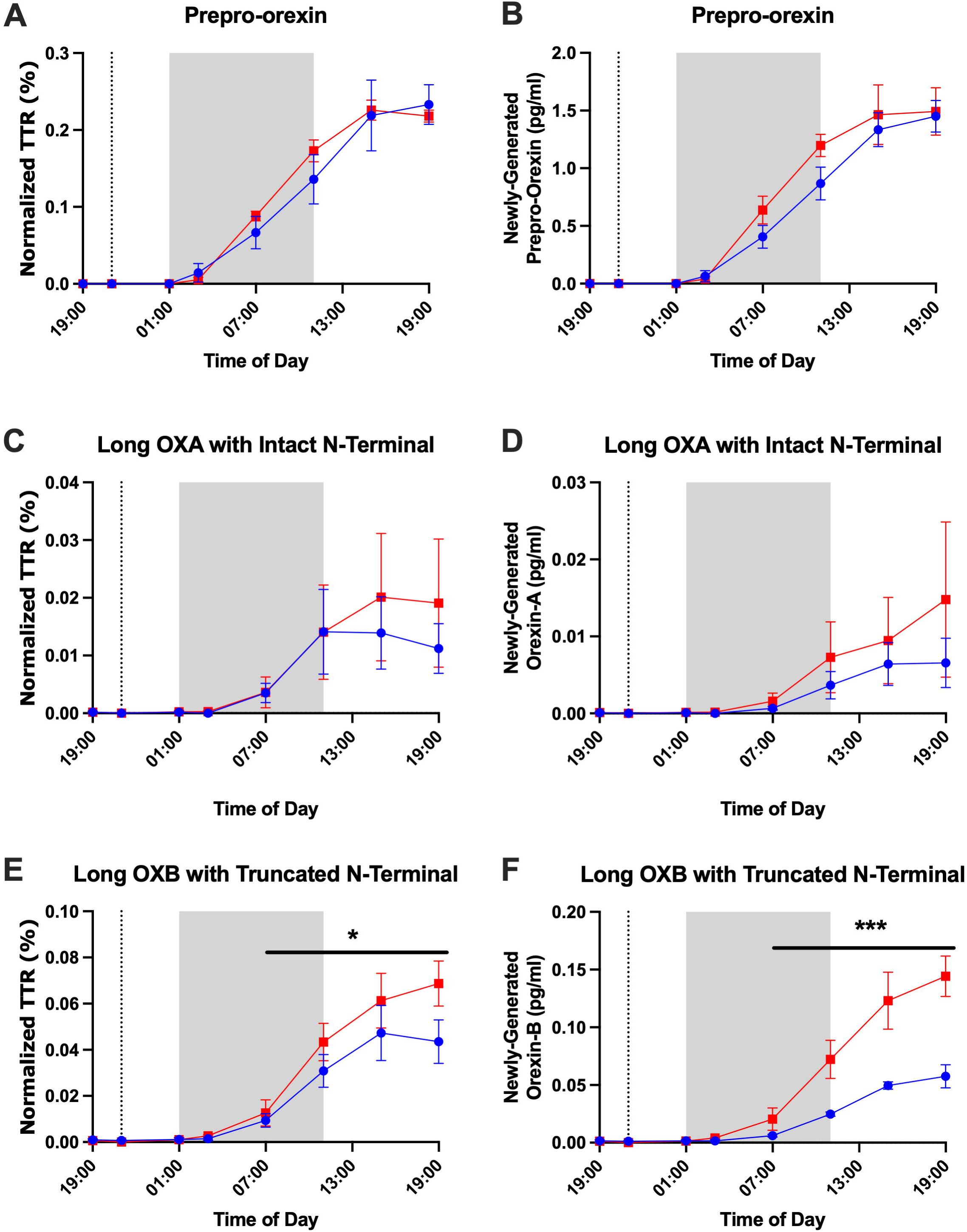
Orexin peptide stable isotope labeling kinetics (SILK). The tracer:tracee ratios (TTR) of prepro-orexin (A), long orexin-A (OXA) with intact N-terminal (B), and long orexin-B (OXB) with truncated N-terminal are shown from 19:00 on Day 1 to 19:00 on Day 2. ^13^C_6_-leucine was infused at 21:00 on Day 1 (vertical dashed line). The shapes of the SILK curves of prepro-orexin are similar for the sleep-deprived and control groups. Orexin-A and orexin-B show lower levels of enrichment in the normal sleep control group compared to sleep-deprived suggesting increased turnover of these peptides in sleep-deprived participants. Red: sleep-deprived. Blue: control. Error bars indicated standard error. Hours 18 to 28 (01:00-11:00) are shaded to account for transit time of cerebrospinal fluid (CSF) from the brain to the lumbar catheter. *p<0.05; ***p<0.001

## DISCUSSION

Orexin peptide dynamics in the human central nervous system are not well-characterized. We report a high-sensitivity mass spectrometry assay that comprehensively measures different forms of prepro-orexin, orexin-A, and orexin-B in human CSF. As expected, all forms of orexin, including the precursor prepro-orexin, were lower in CSF collected from NT1 patients compared to other groups. Previous MS assays found that human CSF orexin-B was undetectable (Lindström et al., 2021), although it has been measured in non-human primates (Narita et al., 2023). We were able to quantify orexin-B peptides and compare across NT1, NT2, and IH. Orexin-B was lower in NT1 participants compared to NT2 and IH. We found that the ratio of short to long metabolites of orexin-B are reduced in participants with NT2 compared to IH, suggesting potential diagnostic utility to differentiate these hypersomnias depending on whether the orexin system disfunction is related or not.

Our MS assay quantified orexin species with long (intact) and short (metabolite) forms of both orexin-A and orexin-B species. Intact orexin peptides are biologically active forms, whereas shorter peptide fragments or truncated metabolites detected in plasma and CSF are hypothesized to represent enzymatic degradation products rather than functional isoforms (Lang et al., 2004). OX1R shows a higher affinity for the highly conserved orexin-A peptide, whereas OX2R demonstrates a more permissive activation profile, responding to orexin peptides with variable amino acid compositions (Ammoun et al., 2003; German et al., 2013). The orexin-A RIA primarily measures short metabolites of orexin-A that are predicted to be inactive metabolites, with <10% of the measured orexin-A coming from intact peptides (Sakai et al., 2019; Vialaret et al., 2025). Shorter or truncated orexin metabolites may have minimal receptor affinity and may represent biochemical indicators of orexin turnover and peptide clearance (Kukkonen, 2021).

In our study with continuous intrathecal CSF sampling during both sleep and wake times, we observed a clear diurnal rhythm of fully processed, long form, mature orexin-A and orexin-B. This supports the role of these biologically active species in the regulation of sleep. Although all orexin peptides were increased during sleep deprivation, we noted differences in how sleep deprivation affected individual orexin peptides. The long form of orexin-A with intact N-terminal, long orexin-B with truncated N-terminal, and short orexin-B with truncated N-terminal were increased from baseline with sleep deprivation compared to the control condition. Since intact orexin-A and orexin-B are biologically active, we hypothesize that the long form of orexin-A with intact N-terminal and the long orexin-B with truncated N-terminal increased due to prolonged wakefulness. The increase in short orexin-B with truncated N-terminal and bimodal increases in short orexin-A with intact N-terminal may represent differences in degradation. Prepro-orexin was increased during sleep deprivation, but this may be due to group differences at the start of the time courses. ^13^C_6_-leucine labeling of orexin peptides found that sleep-deprivation did not affect the labeling of prepro-orexin, but did significantly increase both the TTR and amount of newly-generated peptides for long orexin-B suggesting greater turnover (Bateman et al., 2009; Paterson et al., 2019). Although labeled orexin-A did not significantly differ between groups, the labeling curve increased earlier and peaked higher when participants were sleep deprived. A lack of group differences in orexin-A labeling may be due to high variability resulting from low percent labeling.

In contrast to the findings with labeled long orexin-B, we previously did not observe any turnover differences in the CSF Aβ stable isotope labeling kinetics (SILK) curves in these same participants (Lucey et al., 2018). A potential explanation may be due to the binding of orexin-A and orexin-B to OX1R and OX2R. OX1R and OX2R are G protein-coupled receptors. After activation from binding with their agonists, G protein-coupled receptors typically recruit proteins such as β-arrestin 1 and β-arrestin 2 that promote the internalization of the activated complex into endosomes. Both OX1R and OX2R interact with β-arrestin 1 and β-arrestin 2 (Dalrymple et al., 2011). Internalization of the activated orexin receptor complexes may account for increased turnover as this effectively clears the proteins from the CSF. Prepro-orexin, which neither binds to OX1R or OX2R nor undergoes receptor-mediated internalization, shows no significant change in labeling after sleep deprivation. This suggests that the production and clearance of prepro-orexin remain stable during sleep loss. We hypothesize that the observed increase in orexin-B turnover is driven by the proteolytic cleavage of existing, stored prepro-orexin, rather than by the synthesis of new precursor protein.

A strength of this study is the use of highly sensitive mass spectrometry to reveal new insights into orexin dynamics in human CSF. Previously uncharacterized orexin peptides were measured and we demonstrated the feasibility of quantitating labeled orexin peptides. However, there are several important limitations. Although previously collected longitudinal CSF samples collected over 36 hours under different sleep conditions are a strength, the number of participants is limited. Not all possible labeled orexin peptides could be quantitated and one participant did not have measurable labeled orexins. This study demonstrates the feasibility of the methodology and establishes important avenues for future investigation, including optimizing the ^13^C_6_-leucine labeling protocol. For instance, the relatively low levels of ^13^C_6_-leucine labeling suggests that a greater amount of ^13^C_6_-leucine should be infused for future studies. Longer sampling periods after labeling would also allow for characterization of protein turnover rates and kinetics. Finally, future studies involving greater number of participants are needed to confirm and extend these results. Greater accuracy measuring orexin peptides has the potential to guide the development of pharmacological interventions targeting sleep, such as OX2R agonist for the treatment of NT1 (Dauvilliers et al., 2023), addiction (McKendrick et al., 2025), and other diseases.

## Data Availability

All data produced in the present study are available upon reasonable request to the authors.

## ACKNOWLEDGEMENTS

We thank the participants and families for their contribution to the present study. We thank Akihiko Koyama, Pallavi Sachdev, Jocelyn Cheng, Margaret Moline and Kristin Wildsmith in Eisai for the conceptualization of collaboration between Washington University and Eisai and the discussion to interpret the results.

## CONTRIBUTIONS

K.H. and B.P.L. conceived the study to develop the comprehensive CSF orexin measurements and understand biology. K.H., C.J.S., and R.K.K. executed experiments. C.J.S. and R.K.K. contributed equally to this study. B.P.L. performed statistical analyses. K.H., C.J.S, R.K.K. and B.P.L. analyzed and interpreted the data. G.P. provided the CSF samples and data from NT1, NT2, and IH patients. All authors made contributions to the paper and approved the final version for submission.

