## Supplemental Materials for "Mass Spectrometry-Based Quantification of Orexin Species in Human Cerebrospinal Fluid Reveals Differential Dynamics Associated with Sleep"

| Supplemental Table 1: Mixed Model Results for Longitudinal Orexin Peptides | | | | |
|---|---|---|---|---|
| **Absolute Concentration, pg/ml (23:00-11:00)** | | | | |
| *OXA (RIA)* | | | | |
| Factor | Pairwise Comparison | Mean Difference | F (df) | p-value |
| Intervention | SD vs. C | +26.3 pg/ml | 6.9 (1,32) | **0.013** |
| Time | | | 1.3 (5,32) | 0.297 |
| Intervention x Time | | | 1.9 (5,32) | 0.119 |
| *Prepro-Orexin (MS)* | | | | |
| Factor | Pairwise Comparison | Mean Difference | F (df) | p-value |
| Intervention | SD vs. C | +59.4 pg/ml | 8.5 (1,32) | **0.006** |
| Time | | | 0.84 (5,32) | 0.534 |
| Intervention x Time | | | 0.16 (5,32) | 0.975 |
| *Long OXA with Intact N-Terminal (MS)* | | | | |
| Factor | Pairwise Comparison | Mean Difference | F (df) | p-value |
| Intervention | SD vs. C | +16.2 pg/ml | 24.3 (1,32) | **<0.001** |
| Time | | | 3.01 (5,32) | **0.024** |
| Intervention x Time | | | 0.104 (5,32) | 0.991 |
| *Short OXA with Intact N-Terminal (MS)* | | | | |
| Factor | Pairwise Comparison | Mean Difference | F (df) | p-value |
| Intervention | SD vs. C | +135.2 pg/ml | 16.97 (1,32) | **<0.001** |
| Time | | | 2.2 (5,32) | 0.078 |
| Intervention x Time | | | 2.1 (5,32) | 0.093 |
| *Long OXB with Truncated N-Terminal (MS)* | | | | |
| Factor | Pairwise Comparison | Mean Difference | F (df) | p-value |
| Intervention | SD vs. C | +52.8 pg/ml | 23.9 (1,32) | **<0.001** |
| Time | | | 1.3 (5,32) | 0.281 |
| Intervention x Time | | | 0.29 (5,32) | 0.918 |
| *Short OXB with Truncated N-Terminal (MS)* | | | | |
| Factor | Pairwise Comparison | Mean Difference | F (df) | p-value |
| Intervention | SD vs. C | +20.6 pg/ml | 26.5 (1,32) | **<0.001** |
| Time | | | 1.5 (5,32) | 0.235 |
| Intervention x Time | | | 0.21 (5,32) | 0.957 |
| OXA: orexin-A; OXB: orexin-B; SD: sleep deprived; C: control; MS: mass spectrometry; RIA: radioimmunoassay; pg: picogram; ml: milliliter | | | | |

| **Supplemental Table 2: Mixed Model Results for Longitudinal Orexin Peptides** | | | | |
|---|---|---|---|---|
| **Percent of the Mean (23:00-11:00)** | | | | |
| *OXA (RIA)* | | | | |
| Factor | Pairwise Comparison | Mean Difference | F (df) | p-value |
| Intervention | SD vs. C | +2.7% | 1.3 (1,32) | 0.264 |
| Time | | | 1.9 (5,32) | 0.126 |
| Intervention x Time | | | 2.5 (5,32) | **0.048** |
| *Prepro-Orexin (MS)* | | | | |
| Factor | Pairwise Comparison | Mean Difference | F (df) | p-value |
| Intervention | SD vs. C | +9.6% | 4.5 (1,32) | **0.042** |
| Time | | | 0.72 (5,32) | 0.616 |
| Intervention x Time | | | 0.4 (5,32) | 0.842 |
| *Long OXA with Intact N-Terminal (MS)* | | | | |
| Factor | Pairwise Comparison | Mean Difference | F (df) | p-value |
| Intervention | SD vs. C | +25.8% | 17.7 (1,32) | **<0.001** |
| Time | | | 5.2 (5,32) | **0.001** |
| Intervention x Time | | | 0.22 (5,32) | 0.952 |
| *Short OXA with Intact N-Terminal (MS)* | | | | |
| Factor | Pairwise Comparison | Mean Difference | F (df) | p-value |
| Intervention | SD vs. C | +12.9% | 3.7 (1,32.7) | 0.063 |
| Time | | | 2.03 (5,32.6) | 0.101 |
| Intervention x Time | | | 1.9 (5, 32.6) | 0.126 |
| *Long OXB with Truncated N-Terminal (MS)* | | | | |
| Factor | Pairwise Comparison | Mean Difference | F (df) | p-value |
| Intervention | SD vs. C | +14.7% | 7.4 (1,32) | **0.011** |
| Time | | | 2.7 (5,32) | **0.040** |
| Intervention x Time | | | 0.997 (5,32) | 0.435 |
| *Short OXB with Truncated N-Terminal (MS)* | | | | |
| Factor | Pairwise Comparison | Mean Difference | F (df) | p-value |
| Intervention | SD vs. C | +16.01% | 15.2 (1,32) | **<0.001** |
| Time | | | 2.98 (5,32) | **0.025** |
| Intervention x Time | | | 0.611 (5,32) | 0.692 |
| OXA: orexin-A; OXB: orexin-B; SD: sleep deprived; C: control; MS: mass spectrometry; RIA: radioimmunoassay | | | | |

| Supplemental Table 3: Mixed Model Results for Longitudinal Orexin Peptides | | | | |
|---|---|---|---|---|
| **Orexin Stable Isotope Labeling Kinetics (07:00-19:00)** | | | | |
| *Prepro-Orexin (Normalized TTR%)* | | | | |
| Factor | Pairwise Comparison | Mean Difference | F (df) | p-value |
| Intervention | SD vs. C | +0.102% | 0.414 (1,14) | 0.531 |
| Time | | | 71.66 (3,14) | **<0.001** |
| Intervention x Time | | | 2.602 (3,14) | 0.093 |
| *Long OXA with Intact N-Terminal (Normalized TTR%)* | | | | |
| Factor | Pairwise Comparison | Mean Difference | F (df) | p-value |
| Intervention | SD vs. C | +0.004% | 0.862 (1,14) | 0.369 |
| Time | | | 2.537 (3,14) | 0.099 |
| Intervention x Time | | | 0.296 (3,14) | 0.828 |
| *Long OXB with Truncated N-Terminal (Normalized TTR%)* | | | | |
| Factor | Pairwise Comparison | Mean Difference | F (df) | p-value |
| Intervention | SD vs. C | +0.014% | 8.174 (1,14) | **0.013** |
| Time | | | 19.05 (3,14) | **<0.001** |
| Intervention x Time | | | 0.876 (3,14) | 0.477 |
| **Newly-Generated Orexin Peptides (07:00-19:00)** | | | | |
| *Newly-Generated Prepro-Orexin (MS)* | | | | |
| Factor | Pairwise Comparison | Mean Difference | F (df) | p-value |
| Intervention | SD vs. C | +2.073 pg/ml | 1.97 (1,14) | 0.182 |
| Time | | | 32.94 (3,14) | **<0.001** |
| Intervention x Time | | | 1.042 (3,14) | 0.404 |
| *Newly-Generated Long OXA with Intact N-Terminal (MS)* | | | | |
| Factor | Pairwise Comparison | Mean Difference | F (df) | p-value |
| Intervention | SD vs. C | +0.066 pg/ml | 2.176 (1,14) | 0.162 |
| Time | | | 2.819 (3,14) | 0.077 |
| Intervention x Time | | | 0.293 (3,14) | 0.830 |
| *Newly-Generated Long OXB with Truncated N-Terminal (MS)* | | | | |
| Factor | Pairwise Comparison | Mean Difference | F (df) | p-value |
| Intervention | SD vs. C | +0.967 pg/ml | 34.61 (1,14) | **<0.001** |
| Time | | | 20.62 (3,14) | **<0.001** |
| Intervention x Time | | | 2.786 (3,14) | 0.080 |
| OXA: orexin-A; OXB: orexin-B; SD: sleep deprived; C: control; MS: mass spectrometry; TTR: tracer-tracee ratio; pg: picogram; ml: milliliter | | | | |

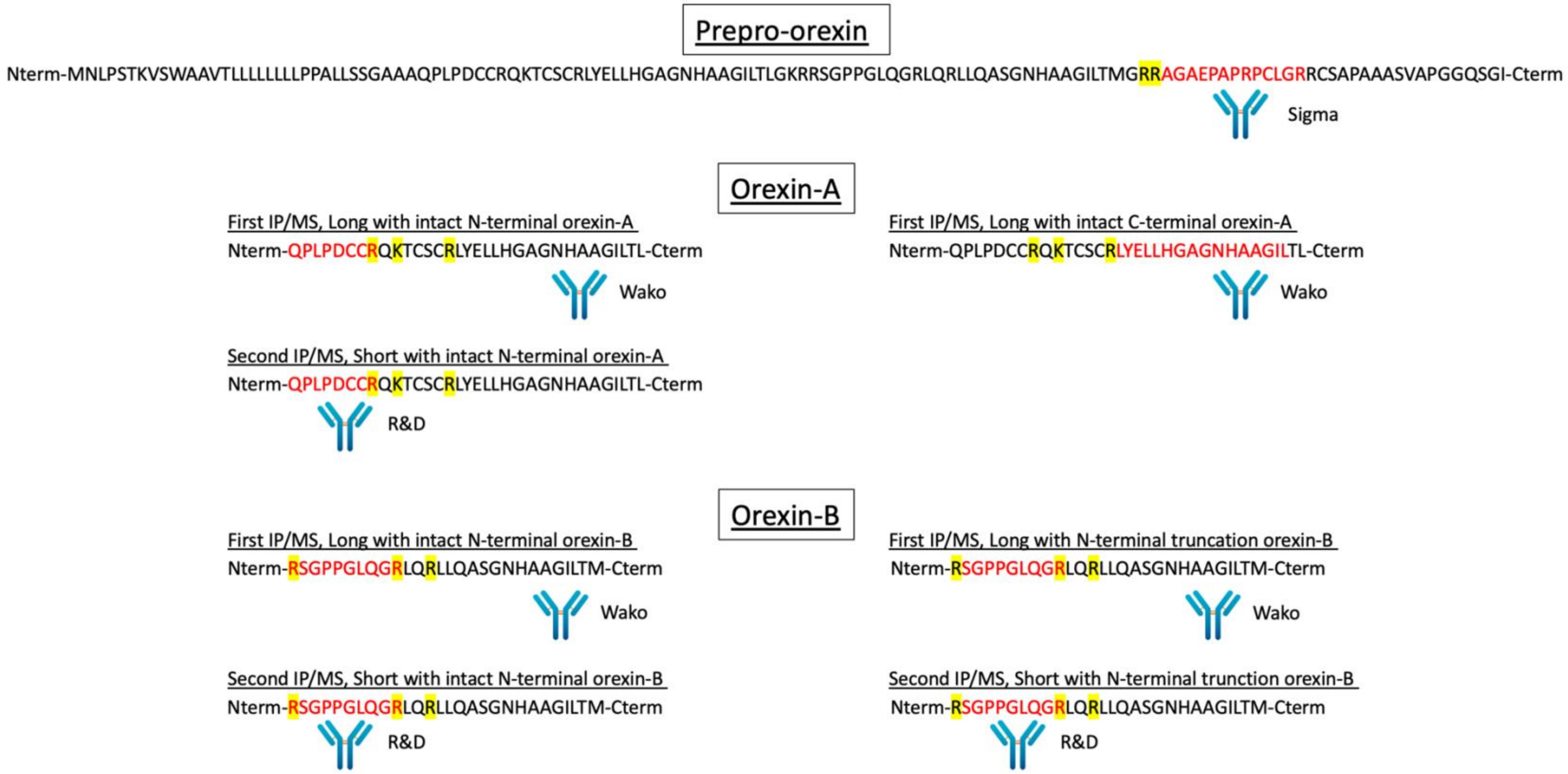

Supplemental Figure 1: Prepro-orexin, orexin-A, and orexin-B peptides measured by mass spectrometry. Yellow: Trypsin cleavage sites. Red: peptides measured by mass spectrometry.

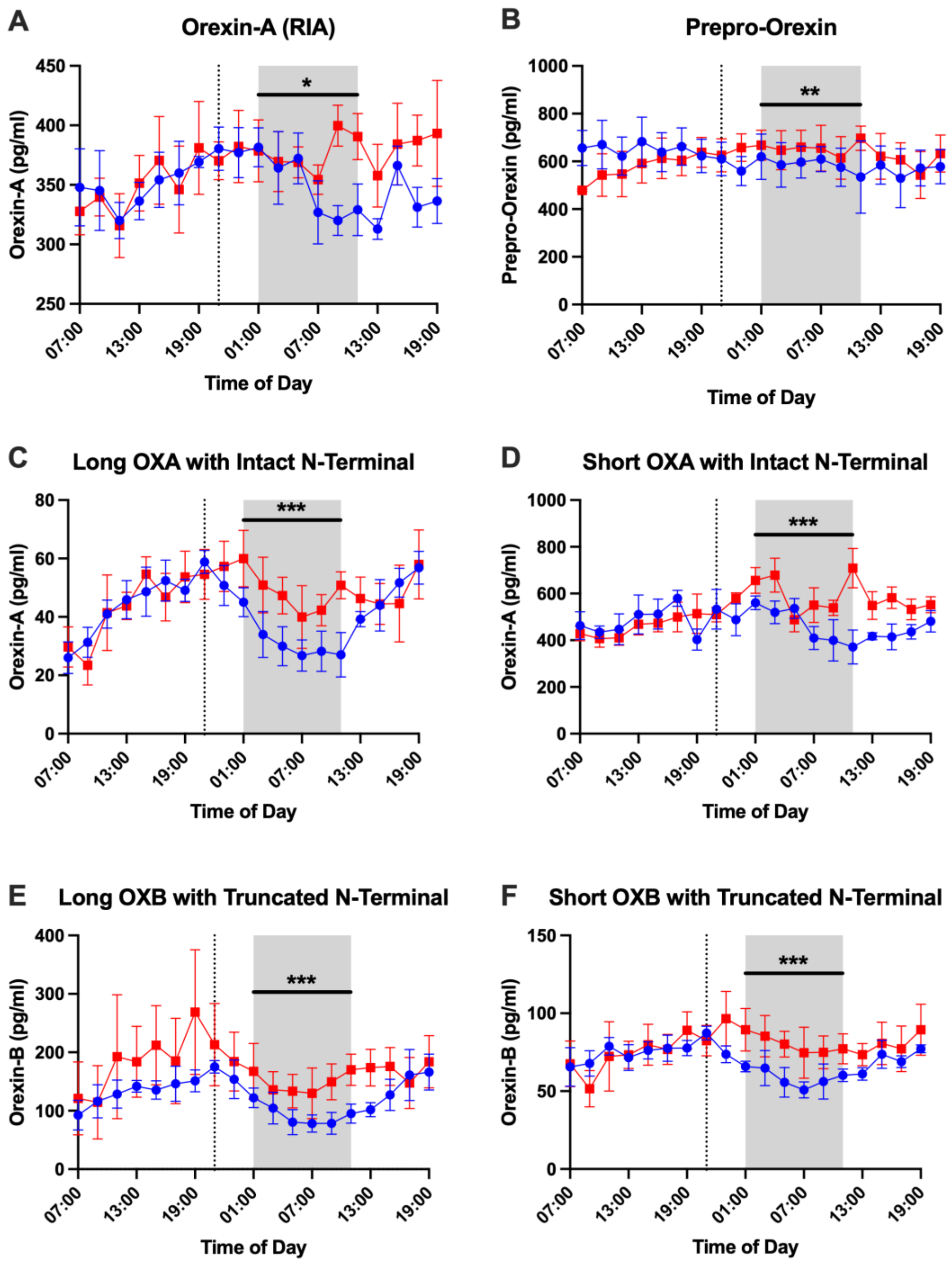

Supplemental Figure 2: Effect of overnight sleep deprivation on orexin peptide concentrations in cerebrospinal fluid. Four participants completed both the control normal sleep and sleep-deprived intervention groups. All orexin peptides are shown in pg/ml concentration. The overnight period during the intervention night was defined as hours 18 to 28 (01:00-11:00) to account for transit time of cerebrospinal fluid (CSF) from the brain to the lumbar catheter (shaded area). Blue: control; Red: sleep-deprived. Error bars indicate standard error. The vertical dashed line is the intervention start time. OXA: orexin-A. OXB: orexin-B. *p<0.05; **p<0.01; ***p<0.001.

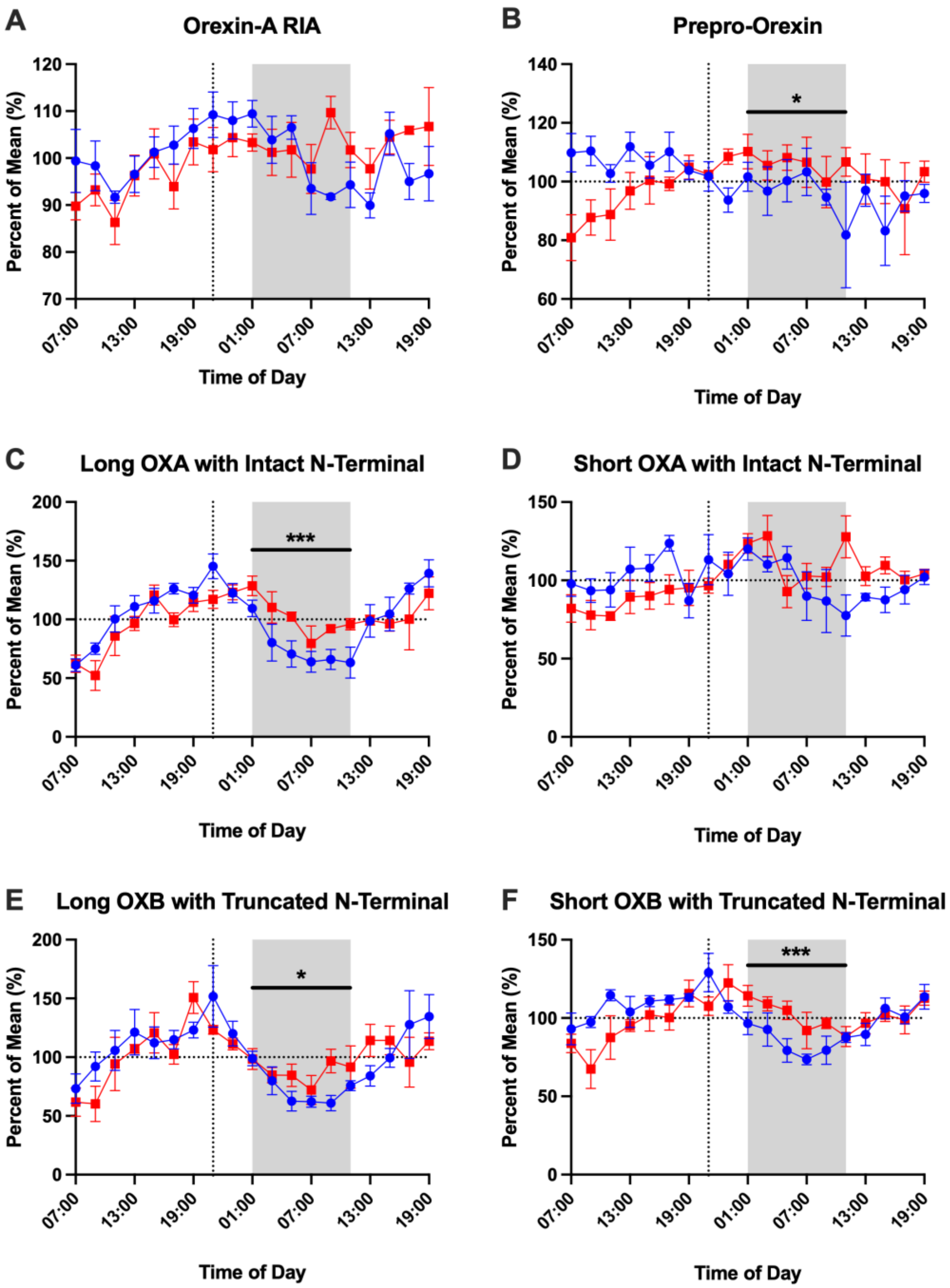

<u>Supplemental Figure 3</u>: Effect of overnight sleep deprivation on orexin peptide concentrations in cerebrospinal fluid normalized to the percent of the mean. Four participants completed both the control normal sleep and sleep-deprived intervention groups. All orexin peptides are shown as percent of the mean of each individual time course. Normalizing to the mean allows for comparing the diurnal oscillation between peptides on the same scale. The overnight period during the intervention night was defined as hours 18 to 28 (01:00-11:00) to account for transit time of cerebrospinal fluid (CSF) from the brain to the lumbar catheter (shaded area). Blue: control; Red: sleep-deprived. Error bars indicate standard error. The vertical dashed line is the intervention start time. OXA: orexin-A. OXB: orexin-B. *p<0.05; ***p<0.001
